# Exploring the lifetime effect of children on wellbeing using two-sample Mendelian randomisation

**DOI:** 10.1101/2022.06.15.22276383

**Authors:** Benjamin Woolf, Hannah Sallis, Marcus R. Munafò

**Author notes:** Corresponding Author: Benjamin Woolf, School of Psychological Science, University of Bristol, Bristol, UK. These authors contributed equally (Hannah Sallis, Marcus R. Munafò).

## Abstract

**Objectives:** To provide Mendelian randomisation evidence of the effect of having children on parental wellbeing.

**Design:** Two sample Mendelian randomisation.

**Setting:** Non-clinical European ancestry participants.

**Participants:** We used the UK Biobank (460,654 male and female European ancestry participants) as a source of genotype-exposure associations, and the Social Science Genetics Consortia (SSGAC) (298,420 male and female European ancestry participants) and Within-Family Consortia (effective sample of 22,656 male and female European ancestry participants) as sources of genotype-outcome associations.

**Interventions:** The lifetime effect of an increase in the genetic liability to having children.

**Primary and secondary outcome measures:** The primary analysis was an inverse variance weighed analyses of subjective wellbeing measured in the 2016 SSGAC GWAS. Secondary outcomes included pleiotropy robust estimators applied in the SSGAC and an analysis using the Within-Family consortia GWAS.

**Results:** The primary IVW estimate found evidence of a 0.153 standard deviation increase for every child a parent has (95% CI: -0.210 to 0.516). Secondary outcomes were generally slightly deflated (e.g. -0.049 [95% CI: -0.533 to 0.435] for the WFC and 0.090 [95% CI: -0.167 to 0.347] for weighted median) implying the presence of some residual confounding and pleiotropy.

**Conclusions:** Contrary to the existing literature, our results are not compatible with a measurable negative effect of number of children on the average wellbeing of a parent over their life course. However, we were unable to explore non-linearities, interactions, or time varying effects.

**Strengths and limitations of this study:**

- Mendelian randomisation (MR) is a natural experiment which is theoretically robust to confounding and reverse causation.
- We were able to use two negative control analyses to explore the robustness of our study to two potential sources of residual confounding (populations structure and passive gene-environment correlation).
- We additionally use pleiotropy robust estimates (like MR-PRESSO, MR-Egger, weighted median, and weighed mode) to explore if our result was affected by direct effects of the genetic variants on the outcome, not mediated by the exposure.
- Because we use summary data, we were unable to explore interactions, non-linear and time-varying, or time sensitive, effects.
- Our study is a proof of concept for using MR to explore the causal effect of the heritable environment.

## Introduction

A well-replicated (1–8), but contested (9–14), finding in the observational quantitative social science literature is a negative association between having children and subjective wellbeing in English speaking countries. Although many studies have failed to replicate the finding in non-English speaking countries (15–17), others have (18,19). For example, Novoa and colleagues found a negative association between having children and subjective wellbeing in Chile (20). Matters are further complicated by non-linearities depending on the age at which wellbeing is assessed in the parents. According to some of the studies having children is not negatively associated with wellbeing when measured in geriatric populations, especially for those with a lower socioeconomic position (SEP), despite a negative association when measured in younger age groups (18,20–23). This is possibly because of increased social support that children can provide to their parents in old age (2,14,21,22).

Because these findings come from mostly cross-sectional, observational studies they should be treated with some caution. For example, most of the studies cited above adjusted for only a few potential confounding variables (e.g., age, sex, and socio-economic position) – if any at all – raising a question of residual confounding. Indeed, Deaton and Stone found that the choice of covariates adjusted for could produce radically different conclusions (15). There will also likely be underlying psychological differences between adults who choose to have children, and those who choose not to have children. Since the examined studies did not adjust for these variables, if these psychological differences also directly influence wellbeing, then they would be an additional source of unmeasured confounding. Relatedly, because most of the literature is derived from correlational surveys, these studies cannot ascertain the direction of effect (6). Finally, cross-sectional studies cannot determine if the apparent change in effect with age is a cohort effect instead, although the corroboration of this finding by some prospective studies makes this less plausible.

Because of the higher risk of bias in traditional observational studies, it is becoming increasingly common in social epidemiology and econometrics to triangulate evidence from conventional observational studies with quasi-experimental designs (24). One such design is Mendelian randomisation (MR) (25–27). In a randomised controlled trial, participants are randomised to an intervention or control arm and followed-up for a certain period. Because genetic variants are inherited at random, comparing outcome status of an individuals with and without a causal variant for the exposure is essentially analogous to a clinical trial (28). In addition, because our genotype is fixed at conception, MR estimates are robust to reverse causation. This also means that any effect estimate derived from MR studies should be interpreted as the lifetime effect of the exposure. Applications of MR have traditionally focused on biomedical exposures where the analogy between MR and randomised controlled trials for a pharmacological intervention is strong because most drugs target proteins, which are the proximal product of genes (29).

MR has been gaining popularity as a method for answering causal questions in psychology and the social sciences in recent years (30,31). Psychiatric genetics and evolutionary theory both suggest that elements of our environment will be genetically influenced, through ‘active gene-environment correlation’ and ‘the extended phenotype respectively’ (32,33). MR has not been used to explore the causal effect of environmental exposures. This is, in part, because it is conceptually less clear how the potential effect of a genetic variant to robustly increase the probability of an environmental exposure – like a traffic accident – by a very small amount is genuinely equivalent to an exposure like being hit by a lorry. Number of children, on the other hand, is a theoretically plausible environmental phenotype for studying with a genetic design like MR because it is the primary endpoint through which evolution by natural selection occurs, and hence should be influenced by genetics (34,35).

Exploring the effect of having children on wellbeing therefore not only answers a question of societal importance, but can also act as a proof of concept for leveraging active gene-environmental correlation within an MR study design to study the causal effect of environmental exposures. We therefore used two-sample MR to explore the lifetime effect of having children on wellbeing.

## Methods

### Study design

We performed a two-sample MR analysis to explore the lifetime effect of having children on wellbeing. Specifically, we used the UK Biobank (UKB) as a source of genetic instruments (36,37), and their weights, for the number of children an individual has, and the 2016 Social Science Genetics Consortium (SSGAC) GWAS meta-analysis of subjective wellbeing as a source of instrument-outcome associations (38).

### Data sources

#### UKB

The UKB is a large (∼500,000 participants) population cohort study in the UK. Members of the public between the ages of 38 and 73, and who lived within 22 miles of an assessment centre, were invited to participate from 2006 to 2010. Approximately 9.2 million individuals were invited to take part, with around 6% participating in the baseline assessment. The sample is 55% female, and predominantly of European ancestry (96%). The full protocol is available online (http://www.ukbiobank.ac.uk/wp-content/uploads/2011/11/UK-Biobank-Protocol.pdf). The study design, participants and quality control (QC) methods have been described in full elsewhere (36). UKB received ethics approval from the North West Multi-Centre Research Ethics Committee (REC reference 11/NW/0382). All participants provided written informed consent to participate in the study. Data from the UKB are fully anonymised.

#### SSGAC

Data on subjective wellbeing was extracted from the 2016 SSGAC subjective wellbeing GWAS (OpenGWAS ID: ieu-a-1009) (38). This was a meta-analysis of 298,420 individuals from 59 studies. Samples included men and women, mostly of European descent living in Europe, North America, or Australia.

#### WFC

Data on wellbeing was also taken from the 2022 WFC sibling estimate GWAS (OpenGWAS ID: ieu-b-4851) (39). This GWAS used the genetic overlap of relatives to adjust for which SNPs were inherited and therefore fully removes most plausible sources of confounding, including ancestry and genetic nurture (40). The consortium combines data on almost 160,000 siblings from 17 cohorts. The GWAS itself had an effective sample size of 22,656 (male and female) individuals of European ancestry. A more detailed description of the individual cohorts included can be found in the paper’s Supplementary Material (39).

### Phenotyping

#### UKB

Information on the number of children (OpenGWAS ID: ieu-b-4760) a participant had, number of full sisters (UKB ID: 1883, OpenGWAS ID: ukb-b-5593), number of full brothers (UKB ID: 1873, OpenGWAS ID: ukb-b-4263), general happiness (UKB ID: 20458, OpenGWAS ID: ukb-b-4062), number of older siblings (UKB ID: 5057, OpenGWAS ID: ukb-b-1997), and hair colour (UKB ID: 1747, Open GWAS IDs: ukb-d-1747_5, ukb-d-1747_4, ukb-d-1747_3, ukb-d-1747_1, ukb-d-1747_2, ukb-d-1747_6) were collected through a questionnaire asked either during the initial visit to an assessment centre or, in the case of general happiness, in an online follow-up. The exact questions asked are provided in the Supplementary Material.

#### SSGAC

This study included measures of life satisfaction, positive affect, or both in the GWAS. The specific questionnaires used to phenotype subjective wellbeing in each sample are described the Supplementary Material (38). These were standardised for the meta-analysis.

#### WFC

All participating cohorts measured wellbeing using a questionnaire. Wellbeing measures were standardised prior to the meta-analysis. More details on phenotyping are provided in the Supplementary Material of the original paper (39).

### Statistical Analysis

#### Overview of the analysis

The primary analysis was an inverse variance weighted meta-analysis of the Wald ratio using independent genome-wide significant SNPs for number of children identified in UKB (37). We additionally used a number of sensitivity analyses, including: a) five pleiotropy robust estimators (MR-Egger, MR-RAPS, MR-PRESSO, weighted median, and weighted mode), b) two sets of negative controls (hair colour, and number of parental siblings) as a falsification test for the presence of residual confounders of the instrument-outcome association, c) using the Within family Consortium (WFC) as a more robust (but less well powered) outcome GWAS, and d) using a less stringent p-value threshold (p<5×10^−6^) for selecting SNPs to increase power. More details can be found in the Supplementary Methods.

#### Assumptions of the analysis

Two-sample MR is an extension of MR to a summary data setting. MR is itself an extension of Instrumental Variables (IV) analysis to genetics. IV makes three assumptions: 1) Relevance: that the variant is robustly associated with the exposure, 2) independence: that there are no variant-outcome confounders. 3) Exclusion restriction: that the variant causes the outcome only through the exposure. For the point estimate to be interpretable, IV analysis additionally has to assume that the instrument-exposure association is monotonic (i.e., if the average causal effect of a genetic variant on number of children is negative, then it is negative for everyone). Finally, two-sample MR makes two additional assumptions: 1) that the GWASs come from the same population. This is required for the MR estimate to be meaningful. 2) That there is no sample overlap. The effect of the second assumption is to make weak instrument bias deflationary (41–43).

Because two-sample MR uses summary data from previously conducted GWASs, if these studies assume, as they typically do, a linear effect, then it is not possible to explore non-linearity using summary data. Traditional IV estimators, such as the Wald ratio used in two-sample MR, can still provide a valid estimate of the average causal effect in the presence of non-linearities (44).

### Sensitivity and additional analyses

#### Negative controls

We used two sets of negative controls to explore two potential sources of confounding. Firstly, hair colour is known to vary by ancestry in the UK/European population, but *prima facie* should not have a direct causal relationship to the number of children we have (45). It can therefore be used as a negative control outcome for population structure (see Figure 1 for a visual representation). Secondly, wellbeing may be affected by our developmental environment. However, the number of children we have is affected by our parental genotype (through inheritance), but this will also influence our developmental environment (through genetic nurture, see Figure 2 for a visual representation). To test if genetic nurture is a residual confounder we used the number of siblings as a negative control outcome, because the number of siblings an individual has is caused by parental genotype, but is unlikely to be caused by the number of children that the individual has. We ran the negative controls only using an IVW estimator because it is the most efficient estimator, so the most likely to detect an association if there is one. In addition, we want an estimator which is not robust to pleiotropy because a pleiotropic association between the genetic instrument and the negative control outcomes (i.e., which is not mediated by number of children) is still evidence of an association between the instrument and a confounder of the instrument-outcome association, and hence a violation of the independence assumption.

**Figure 1:**
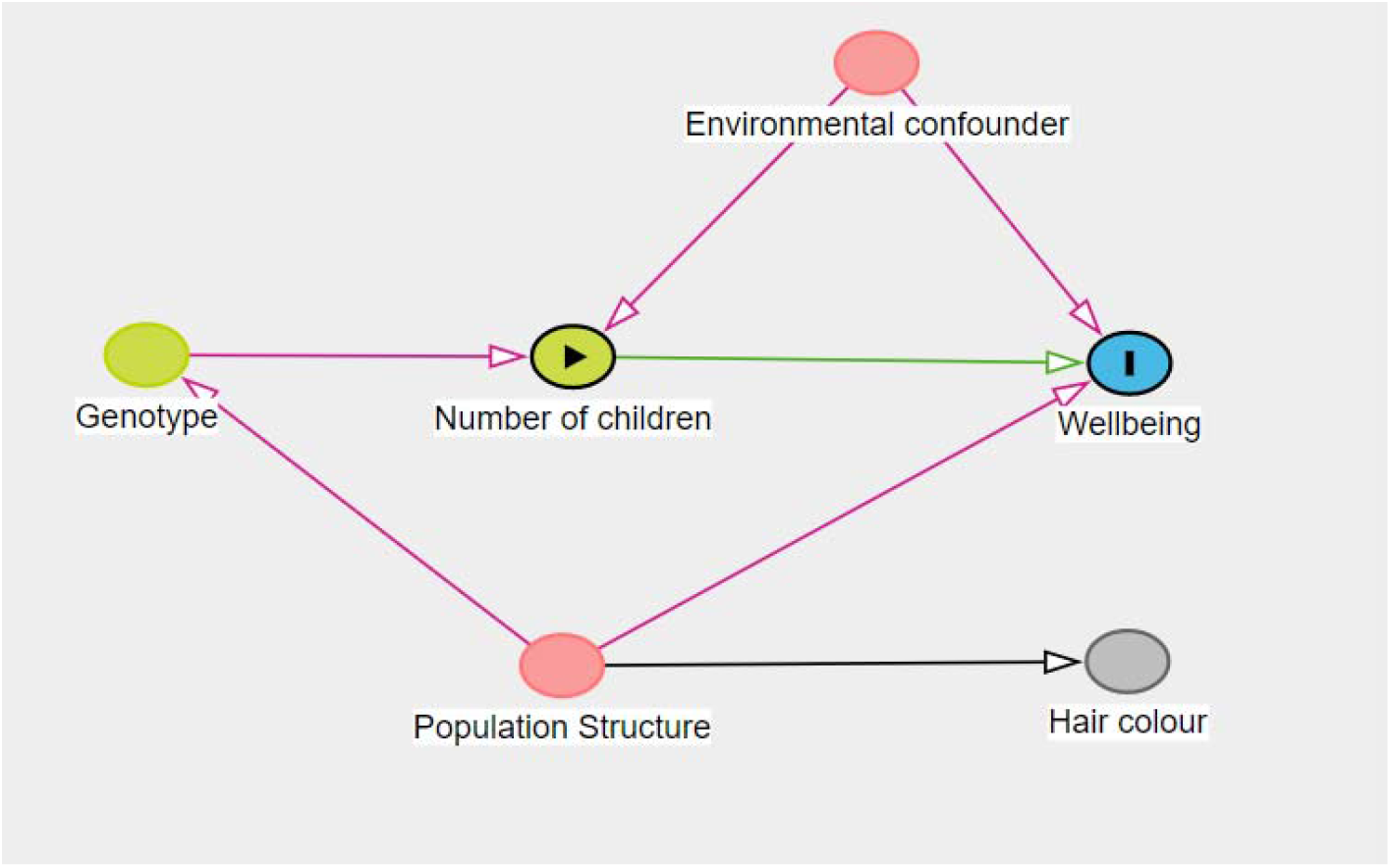
Directed Acyclic Graph for the hair colour negative control outcome. Hair colour is hypothesised to associated with population structure, but to not have a direct causal link to either the genetic instruments or wellbeing. Therefore, any association between the genetic instruments and hair colour will be due to a residual confounding effect of population structure.

**Figure 2:**
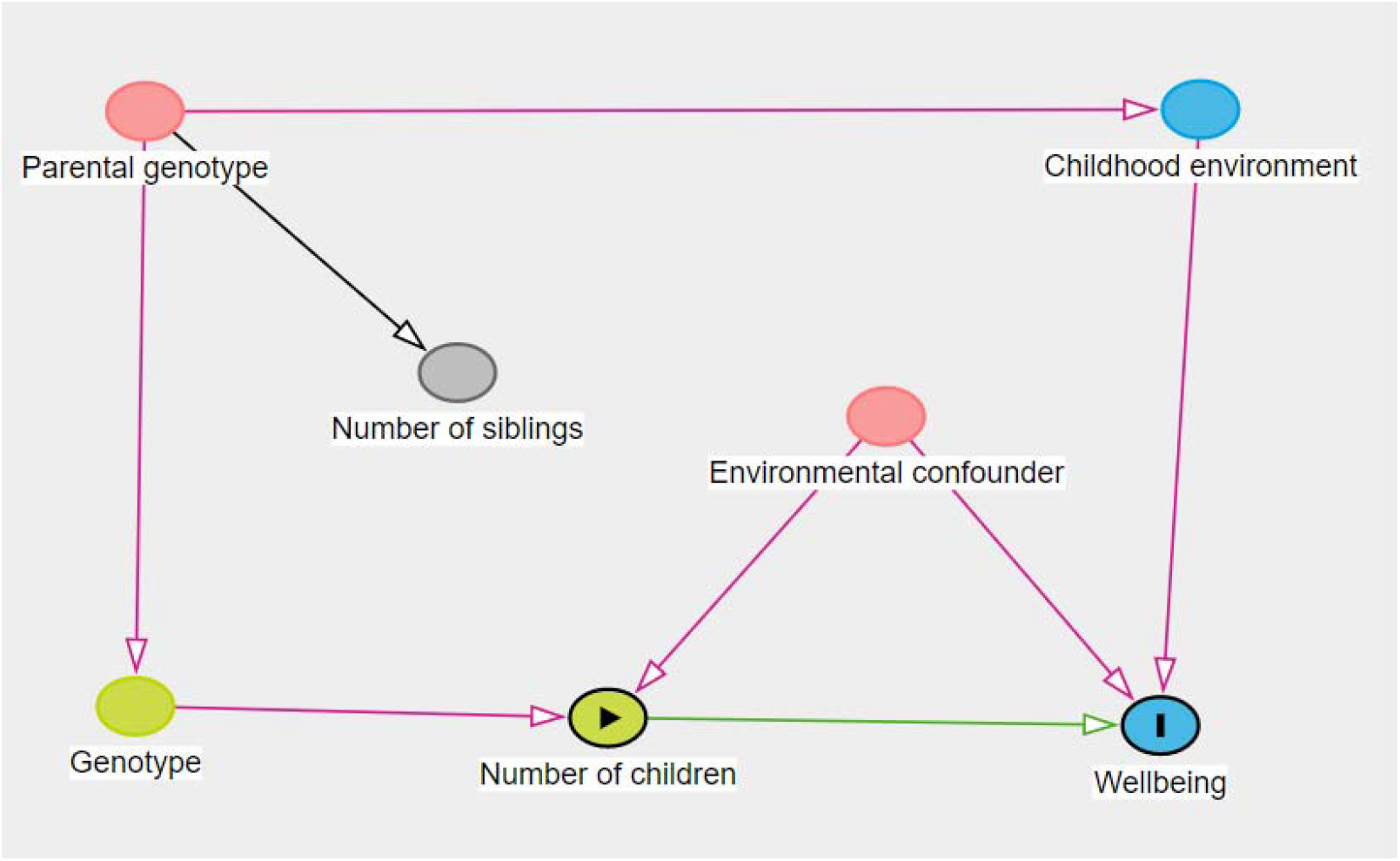
Directed Acyclic Graph for the number of siblings negative control outcome. The parental genotype determines not only their children’s genotype but also, via the parental phenotype, the environment in which the children grow up. If someone’s childhood environment influences their wellbeing latter in life, then the parental genotype would confound any association between an individual’s genetic instruments and wellbeing. Because, in the UK, parents typically stop having new children before their children start having children, it unlikely that a child having children will influence his or her parents to have more children. Hence, the number of siblings an individual has should not be caused by an individual’s genetic liability to having children, but will be influenced by the parental liability to having children. Therefore, any association between an individual’s genetic liability to having children and the number of siblings they have would be an indicator of residual confounding due to the parental genotype.

#### WFC GWAS

Family data has been proposed as a way of eliminating the risk of confounding in MR studies (40). Potential violations of the independence assumption, like population structure and genetic nurture, occur because the distribution of SNPs in a population GWAS is only approximately random. By conditioning on the parental genotype, however, the back door path used by these biases is blocked. Although these GWASs are less biased, the use of non-independent observations means that they need much larger samples than a population GWAS to achieve the same level of power. We, therefore, used the WFC GWAS of well-being as an additional sensitivity analysis to explore the robustness to potential confounders.

#### Less stringent SNP selection

Power in two-sample MR studies is a function of instrument strength, the precision of the outcome GWAS, and the number of instruments. Since we are limited to using pre-collected data, the outcome GWAS’s precision cannot be varied. However, a p-value threshold of 5 × 10^−6^ equates to an F-statistic of approximately 10, and should therefore not lead to weak instruments, but because it is 100-fold larger, should increase the number of SNPs used in the analysis. We therefore also used SNPs with an indicative association (p < 5 × 10^−6^) with the exposure to explore how sensitive the primary analysis was to a potentially better-powered set of instruments.

#### Leave-one-out analysis

We additionally used a ‘leave-one-out’ analysis and the MR-PRESSO outlier test, as part of our MR-PRESSO analysis, to explore if any of the variants were outliers and had a disproportional effect on the overall IVW estimate. This sensitivity analysis works by excluding each SNP in turn and running the IVW analysis without the excluded SNP.

Additional information, including details on genotyping, instrument construction, and MR estimators can be found in the supplementary methods.

## Results

### Descriptive data

#### Number of participants and SNPs in each sage

The UKB exposure GWAS had information from over 460,00 participants on almost 10 million SNPs. The primary outcome analysis used information on six of these SNPs which were genome-wide significant for number of children from 298,420 participants from the SSGAC. This was increased to 50 SNPs by using a 5 × 10^−6^ p-value threshold. The WFC outcome data used information on 8 genome-wide significant SNPs with an effective sample size of 22,656 participants (Figure 3).

**Figure 3:**
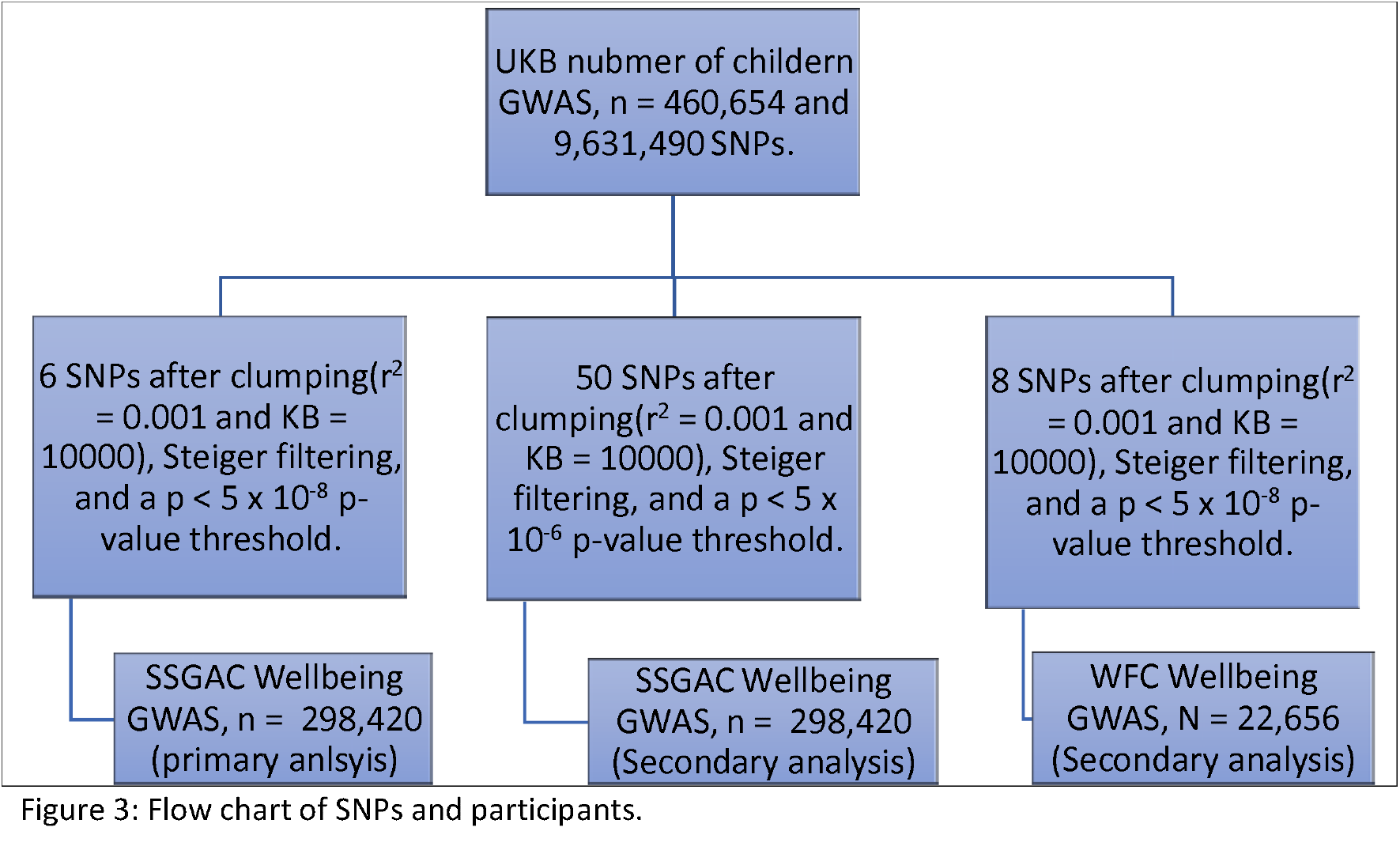
Flow chart of SNPs and participants.

#### Two-sample MR specific assumptions

Both outcome samples had some overlap with the UKB. The SSGAC does not state how many UKB participants were included, however around 157,000 UKB participants provided information on the measure of general happiness used by the SSGAC, which entails a maximum sample overlap of around 53% for the SSGAC and 34% for the UKB. The UKB also contributed around 4,250 sibships to the WFC wellbeing GWAS, which equates to around 19% of the WFC sample and 2% of the UKB sample.

Because of all GWASs were drawn from European populations of both males and females, with some overlapping participants, it seems likely that the samples can all be treated as coming from the same population. We also found no evidence of a difference in SNP effect estimates for the SNP-outcome associations in the UKB and the WFC or the SSGAC consortium using either a 5×10^−6^ p-value or 5×10^−8^ p-value threshold which further supports this conclusion (Supplementary Figures 1-3).

### Main results

The primary IVW estimate indicates a 0.153 (95% CI -0.209 to 0.515) standard deviation change in wellbeing for every additional child an individual has (Figure 4 and Supplementary Figure 4). In most of the samples used in the SSGAC GWAS a 1 to 2 standard deviation change is equivalent to a 1 unit increase on a 5-level psychometric question. For example, the 23andMe study asked participants to rate from very dissatisfied with their life (score = 0) to very satisfied with it (score = 5) and had a standard deviation of 1. This means that a one standard deviation increase would be the same as going from very dissatisfied to somewhat dissatisfied. Supplementary Tables 1 to 3 provide the gene-exposure and gene-outcome associations used in this study.

**Figure 4:**
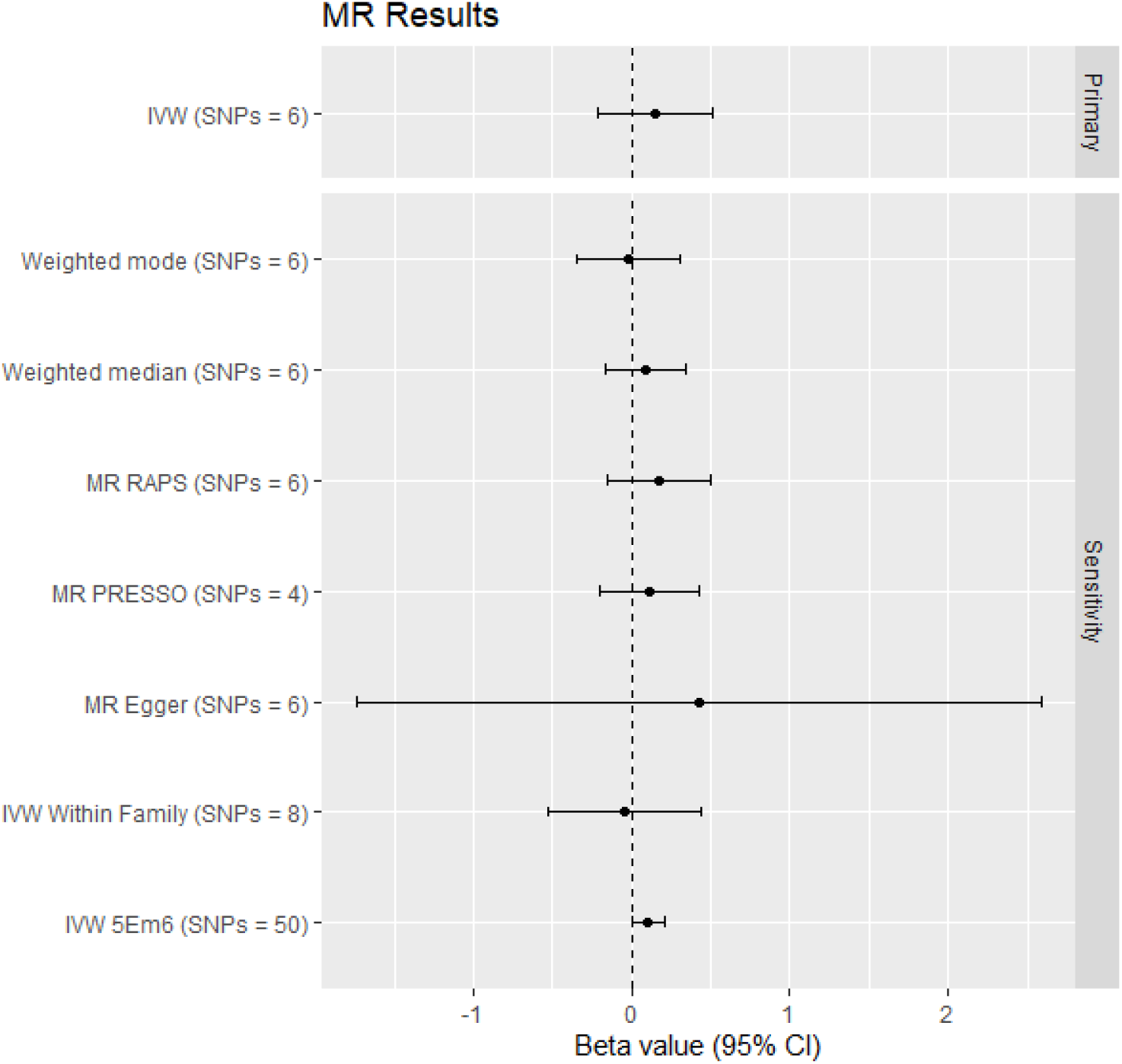
Forest plot of the primary and sensitivity analyses

### Assessment of assumptions

#### Weak instrument bias and NOME

For the primary analysis, the F statistic was 49, and the I^2^ for the instrument-exposure association was 98%. These both imply that there would be around a 2% error in the MR estimates due to weak instrument bias. For the analysis using the WFC, the F statistic was 44, and the I^2^ 98%. For the analysis using a less stringent p-value, the F statistic was 25, and the I^2^ 96%.

#### Heterogeneity and exclusion restriction violations

The Cochrane Q statistic for the Wald ratios of the primary analysis was 24.59 (p < 0.001) and the I^2^ for the Wald ratio was 80%. Together with the asymmetric funnel plot (Supplementary Figure 5), and the MR-PRESSO global test for outliers (p = 0.006) this implies the presence of some pleiotropic SNPs. However, the Egger intercept was -0.004 (SE = 0.017, p = 0.815). Similar results were found for the secondary analyses (Supplementary Table 4).

### Sensitivity and additional analyses

#### Pleiotropy robust estimators

The pleiotropy robust estimates were mostly similar to the IVW estimate, although generally slightly deflated (Figure 4 & Supplementary Table 4). The exception to this was the MR-Egger estimate, although the wide 95% confidence interval (which overlaps with the IVW estimate) implies that this could be due to a lack of precision. In addition, the MR-RAPS estimate was slightly inflated, probably because RAPS is robust to both moderate amounts of weak instrument bias and pleiotropy.

#### Negative controls

The negative control outcome analysis did not find any evidence of an association of the instruments with hair colour, however, there was evidence of an association with two out of three of the sibling questions (p < 0.001 for number of full brothers and p = 0.005 for number of full sisters) implying the possibility of some residual confounding (Supplementary Table 5).

#### WFC outcome

Consistent with the negative control analysis, the WFC secondary analysis showed deflated point estimates compared to when using the SSGAC outcome GWAS (Supplementary Table 3). For example, the IVW estimate is -0.049 (95% CI: -0.533 to 0.0436).

#### Less stringent SNP selection

The standard error of the IVW estimate when using a 5×10^−6^ threshold was more than three times smaller than when using the more traditional 5×10^−8^ threshold (Supplementary Table 4).

#### Leave one out analysis and MR-PRESSO outlier test

The MR-PRESSO outlier test for the primary analysis identified rs10270358 and rs72687493 as outliers. An exploratory search of Phenoscanner showed no phenotypes associated with rs72687493 but found that rs10270358 is associated with seeing a doctor for anxiety or depression as well as chronic disability/infirmity both of which could reduce wellbeing (46). However, these SNPs did not seem to be introducing a bias in the leave-one-out analysis (Supplementary Figure 6) and the outlier test did not detect any outliers in the secondary analyses.

### Pre-specified interpretation

#### Pleiotropy

Because the indicators for the presence of pleiotropy (like the I^2^ and Cochrane Q statistics for the Wald ratio, funnel plot and MR-RAPS) all indicated the presence of pleiotropy, and because the ‘pleiotropy robust’ estimators generally had deflated estimates compared to the IVW estimate, it seems likely that the IVW estimates are being inflated by some residual pleiotropy.

#### Residual confounding

The association of the instruments with two out of three of the sibling negative controls combined with the change in estimate from the WFC GWAS implies that there was some inflation due to residual confounding from genetic nurture in the primary IVW estimate.

#### Low Power

The number of SNPs increased almost nine-fold in this secondary analysis when compared to the primary one. This resulted in a three-fold decrease in the size of the standard error (0.185 to 0.053) for the IVW estimate, and implies that there is also a large amount of residual random error in the estimates. However, the point estimates in this analysis were generally deflated when compared to the primary analysis. This should be in part explained by the approximate halving of the F statistic.

## Discussion

The existing observational literature implies that having children is detrimental to parental wellbeing. However, our primary analysis found a positive 0.153 (95% CI -0.209 to 0.515) standard deviation change in wellbeing for every additional child an individual has. If we assume the measures are on a ratio scale, that people score the nearest category to what they feel, and that a one SD increase welling is a unit increase in on a 5-level psychometric question, then our results would be compatible with a one-unit increase on this scale for each child someone has (e.g., from neither satisfied nor dissatisfied to somewhat satisfied), but incompatible with a measurable negative change in subjective wellbeing.

However, our additional and sensitivity analyses imply that this may overestimate the true effect. Our negative control analysis found that our instrument was associated with the number of siblings, and that the point estimate was deflated when using the WFC outcome GWAS, implying the presence of residual confounding due to genetic nurture. Likewise, the heterogeneity statistics implied the presence of residual confounding, while most pleiotropy robust estimators were again deflated. It is therefore likely that the true effect will be smaller than a measurable increase (i.e. a one unit increase in on a 5-level psychometric question) in well-being for every child a parent has.

### Generalisability

One possible explanation for the discrepancy between the observational and MR estimates is that the target estimands are not directly comparable. If we assume that age does not modify the variant-exposure association, then MR estimates should be interpreted as the average effect of the exposure on an outcome over the entire lifetime (47). This means that transient effects of having children on wellbeing, such as the stress of looking after a newborn baby, will not be detectable in a typical MR design. Some studies found that having children was beneficial to parental wellbeing in old age (18,20–23). One possible explanation for the discrepancy between our, and the existing literature’s, results would therefore be that the transient negative effect is counterbalanced by the latter positive ones –resulting in an average effect close to zero. Although methods for addressing time-varying exposures are currently being developed (48), there is still no consensus on how to best deal with transient effects within an MR framework. We are therefore unable to empirically explore this interpretation further.

Additionally, our estimates were all drawn from European samples. Because some of the existing literature had found different effects in, predominantly non-European, non-English speaking populations to those observed in English speaking populations (15–17), our results may not generalise to other populations.

### Strengths and limitation

This study has several methodological strengths. Firstly, we believe it is the first study to apply MR to explore the effects of the heritable environment in a setting in which gene-environment equivalence is plausible. By doing so, we have been able to leverage the methodological strengths of MR, such as improved robustness to confounding and reverse causation to explore the effect of having children on wellbeing. Secondly, we believe that this is the first study to have explored the ‘same-population’ two-sample MR specific assumption by testing for a difference in the SNP effects. This test is motivated by the intuition that effects drawn from the same population should only differ because of chance. We are unaware of any existing quantitate tests of this assumption, and therefore hope that it will be useful in further applied MR studies.

There are also methodological limitations to this application of two-sample MR. As already noted, we were unable to explore time-sensitive effects. Relatedly, we were forced to assume a linear dose-response for the effect of the number of children on wellbeing. We considered a sensitivity analysis using individual-level data, but ultimately decided against doing so due to a lack of sufficiently good individual-level data: Of the two available data sources, ALSPAC had detailed phenotyping, but on a relatively small number of participants (∼2,000). Because non-linear MR is less well powered than a linear MR, this sample would therefore be underpowered for this analysis. On the other hand, the UKB, only had a five-level minimal phenotype for happiness. Since poor phenotyping can mask non-linearities, and because happiness may not be the same as wellbeing, the interpretation of any analysis in the UKB would be unclear (49). MR is generally more robust as a test of the causal null hypothesis than as a method of effect estimation because of many of the complications, like those described above, of interpreting MR effect estimates. However, this approach to interpreting MR results may be less robust here because our sensitivity analyses implied that our study may be underpowered.

### Summary and conclusions

We conducted a two-sample Mendelian randomisation study to explore the causal effect of having children on wellbeing. Contrary to the previous literature, our results imply the presence of a small-but-meaningful positive lifetime effect of having children, although this may be due to violations of the exclusion restriction and independence assumptions. Comparing our results to the existing observational studies is complicated by the temporal insensitivity of MR estimates. Future studies could therefore consider using other quasi-experimental methods, such as Interrupted Time Series (50), to explore if the discrepancy between our findings and the observational literature are due to transient effects of having children which MR is either unable to detect or which average out over the life course.

## Data Availability

All data that was used in this study is publicly available from the MRC-IEU OpenGWAS platform using the IDs provided in the text.

## Other Information

### Funding

Benjamin Woolf is funded by an Economic and Social Research Council (ESRC) South West Doctoral Training Partnership (SWDTP) 1+3 PhD Studentship Award (ES/P000630/1). BW, HS and MM work in the MRC Integrative Epidemiology Unit which is supported by the University of Bristol and UK Medical Research Council (MC_UU_00011/1, MC_UU_00011/3, MC_UU_00011/7). This research received no specific grant from any funding agency in the public, commercial or not-for-profit sectors.

### Data and data sharing

All data that was used in this study is publicly available from the MRC-IEU OpenGWAS platform using the IDs provided in the text. The R code used in the manuscript is available from https://doi.org/10.17605/OSF.IO/BTPH9

### Conflicts of interests

The authors declare no conflicts of interest.

## Notes

### Competing Interest Statement

The authors have declared no competing interest.

