## Supplementary Methods and Figures for "Exploring the lifetime effect of children on wellbeing using two-sample Mendelian randomisation"

Questions asked in the UKB

Information on siblings was ascertained through three questions asked at baseline recruitment. Number of full sisters (UKB ID: 1883, OpenGWAS ID: ukb-b-5593) asked the question "How many sisters do you have? (Please include those who have died, and twin sisters. Do not include half-sisters, step-sisters or adopted sisters)". Number of full brothers (UKB ID: 1873, OpenGWAS ID: ukb-b-4263) asked “How many brothers do you have? (Please include those who have died, and twin brothers. Do not include half-brothers, step-brothers or adopted brothers)"; and the number of older siblings (UKB ID: 5057, OpenGWAS ID: ukb-b-1997)  "How many OLDER brothers/sisters do you have? (Please include those who have died, and twins. Do not include half-, step- or adopted brothers and sisters)". Hair colour (UKB ID: 1747, Open GWAS IDs: ukb-d-1747_5, ukb-d-1747_4, ukb-d-1747_3, ukb-d-1747_1, ukb-d-1747_2, ukb-d-1747_6) was ascertained through a questionnaire asked at baseline assessment. Participants were asked: "What best describes your natural hair colour? (If your hair colour is grey, the colour before you went grey)". To measure general happiness (UKB ID: 20458, OpenGWAS ID: ukb-b-4062) participants were ask "In general how happy are you?" and could select options from “Extremely happy”, “very happy”, “moderately happy”, “moderately unhappy”, “very unhappy”, or “extremely unhappy”.

Genotyping

*UKB:* All UKB GWASs which were used in this study were conducted using the MRC-IEU UKB GWAS pipeline (1). A full description of the pipeline methods can be found elsewhere. In brief, after a standardised quality control process, and imputation using the UK10K Haplotype Reference Consortium, the summary statistics were created using a linear mixed model implemented in BOLT-LMM (2), adjusting for sex and SNP-chip. Further information about each GWAS, including the number of participants included for each measure, can be found at <https://gwas.mrcieu.ac.uk/>.

*SSGAC:* Genotyping and QC information about the included samples are described the Supplementary Material original publication (3). The participating cohorts were asked to adjust their GWASs for the first four principal components of the genetic relationship matrix, as well as sex, age, age squared, and study-specific covariates for batch/study cite effects where appropriate. The LDSC intercept for the GWAS is 1.0188 (SE = 0.0076) implying the presence of a very small amount of residual population structure.

*WFC:* The participating cohorts from the WFC ran GWASs using a within-sibship model. This model adjusts for the mean genotype of each participant’s sibling, and inflates the standard error to account for clustering. The model additionally adjusted for age, sex, and the first twenty principal components. The results of the individual studies were then meta-analysed using a fixed-effects model. More details on the GWAS methods, including the QC employed, can be found in the original publication (4). The LDSC intercept for the GWAS implied no evidence of residual population structure (b = 1.005, SE = 0.0063).

Instrument construction

Genetic instruments were selected using a statistical criterion of having a genome-wide significant association with the exposure (p < 5 x 10^-8^). We additionally clumped the variants using an r^2^ of 0.001 and KB of 10,000, thereby ensuring that the instruments are independent of each other. Genetic variants which have their gene-exposure association estimated in the same dataset used to select instruments can suffer from a bias called ‘Winner’s Curse’. This occurs because variants can meet the statistical criteria due to having a genuine association or because they, by chance, have an unusually large amount of noise. This results in variants appearing to have a larger association than they actually do. We therefore further filtered these variants using the False discovery rate Inverse Quantile Transformation (FIQT) winners curse correction developed by the SSGAC (5). This uses an analogy between multiple testing and winners curse to apply an easy to implement correction to effect estimates.

We used the TwoSampleMR R package to harmonise the two GWASs. Palindromic SNPs were only excluded if their allele frequency could not be used to infer which strand was positive. In cases where SNPs are missing in the outcome dataset, we used TwoSampleMR to automatically impute LD proxy variants, using an r^2 of 0.8 from the European subsample of the 1000 genomes project.

MR assumes that the causal pathways go from the genetic variant to the exposure to the outcome. However, when selecting genetic instruments using a data-driven method, as we did, it is possible to select some SNPs where the causal path is from the variant to the outcome to the exposure. When the exposure and outcome GWASs are of similar power, one should expect SNPs to explain a larger proportion of the variance in the more proximal GWAS than the more distal one. Steiger filtering, which we applied, uses this logic as a method of removing SNPs that are more proximal to the outcome than the exposure.

Statistical methods

The primary MR estimator in this study was the Wald ratio. This is defined by the variant-outcome association divided by the variant-exposure association. Because both the variant-exposure and variant-outcome associations were derived from linear models, MR analysis also assumes a linear model.

We then used six methods of meta-analysing the MR estimates for each SNP: IVW, MR-Egger, weighted median, weighted mode, MR-RAPS, and MR-PRESSO. The IVW estimate will return the true effect if all the IV assumptions are valid. The other five other, ‘pleiotropy robust’, methods can return the true effect if some of the instruments are invalid, but have reduced power. Specifically, the weighted mode assumes that the modal effect size is valid estimates of the true effect size, while weighted median assumes that at least half of the SNPs are valid. IVW can be thought of as a regression of the variant-outcome association on the variant-exposure association, with the intercept fixed at zero (6). MR-Egger extends this model to allow for a non-zero intercept. If we assume that the variant-exposure effect size is independent of the size of any bias (such as a pleiotropic effect) then the biasing pathways should impact the intercept but not the slope. This assumption is called the INSIDE assumption. Additionally, MR-Egger assumes that there is no measurement error in the exposure GWAS (called the NOME assumption) (7,8). MR-RAPS assumes that pleiotropic SNPs are outliers (modelled by a random effects parameter with a mean of zero) and therefore down weights them in a random effect meta-analysis. We implemented MR-RAPS using a square error loss function, accounting for overdispersion (i.e., systematic pleiotropy) (9). MR-RAPS is robust to both balanced pleiotropy in non-outlier SNPs and weak instrument bias. MR-PRESSO first runs a global test for pleiotropy, and then tests for outliers. It then excludes outliers before running a meta-analysis and testing for a difference between the MR estimates with and without the outliers. MR-PRESSO assumes INSIDE and that >50% of SNPs are valid instruments (10).

Assessment of assumptions

Weak instrument bias is inversely proportional to the F-statistic of the variant-exposure association, with a common threshold of ten being required. We therefore calculated the mean F statistic for the variant-exposure association, as well as the F-statistic for each SNP. MR assumes that there are no confounders of the variant-outcome association.

Although the other assumptions made in an MR analysis are not provable, it is possible to run falsification tests on many of them. To ensure that population structure was controlled for we checked that the GWASs used either BOLT-LMM, which adjust the GWAS for the entire genetic relationship matrix (2), or adjusted for principal components of the genetic relationship matrix. LD score regression (LDSC) can be used to explore residual population structure in GWASs that did not use linear mixed models like BOLT-LMM. Specifically, if the LDSC intercept is very different from one, this implies the presence of residual population structure (11). We also ran a set of sensitivity analysis, described below, as falsification tests for residual gene-outcome confounders.

Horizontal pleiotropy occurs when a genetic variant associates with two phenotypes for independent reasons. This can cause a violation of the exclusion restriction assumption. However, if horizontal pleiotropy is present, the exact pathway should be different for each variant. It has therefore been argued that, if it occurs, it should create heterogeneity in the MR estimates, and its presence can therefore be tested using a heterogeneity statistic. The presence of horizontal pleiotropy was also visually explored using a funnel plot.

The NOME assumption can be tested by checking that the I^2^ statistic for the gene-exposure association is greater than 90% (7).

We checked sample overlap by checking which samples were reported as being included in each of the GWAS consortia in the respective publications. Two-sample MR studies generally validate the assumption that samples are drawn from the same population by checking that the studies are demographically similar (12). However, in situations where there instrument-exposure or instrument-outcome association has been measured in both the exposure and outcome samples it can be possible to validate this assumption quantitively. One would expect to find only chance differences in estimates drawn from the same population (13,14). Therefore, on top of comparing demographic information, because information on happiness from the UKB was used in the SSGAC GWAS, we checked that the average difference SNP estimates from the UKB compared to the SSGAC and WFC were approximately zero. Because the differences in the precision of the measure of wellbeing between the UKB and SSGAC could introduce heterogeneity into the estimation of the difference in SNP effects, we chose a random effects meta-analysis as the primary estimator.

*Triangulation and planned interpretation of sensitivity analyses and negative controls*.

*Pleiotropy*. Pleiotropy can bias MR estimates in either direction. However, on the assumption that pleiotropy introduces the statistical properties described above, if it is present we would expect to see evidence of heterogeneity in the SNP effect, and possibly outliers in the leave-one-out analysis. The different sensitivity analyses are robust to different types of pleiotropy and therefore can behave differently from each other. For example, if the MR-Egger and/or MR-PRESSO sensitivity analyses produce very different estimates from the other sensitivity analyses then this could be an indication of a violation of the InSIDE assumption. We would therefore interpret the sensitivity analyses as indicating the presence of pleiotropy if either all produce very different estimates from each other, or a plurality of them produce a consistently different estimate from IVW as an indication of residual pleiotropy.

*Residual confounding*. If there is residual confounding due to population structure or passive gene-environment correlation, then we would expect to find consistent evidence in their respective negative control analysis. Because residual confounding can also bias MR estimates in either direction, we would then expect this to lead to the point estimates between the SSGAC and WFC GWAS to be different, although a small chance variation is to be expected because of the reduced precision in the within family GWAS.

*Low power*. If the power of the study can be improved by adding more weakly associated SNPs then we would expect that the 95% CI for the IVW estimate would be smaller when using a p < 5 x 10^-6^ threshold than the genome-wide significant one. However, interpreting any change in the point estimate after this sensitivity analysis can be difficult because the use of weaker instruments in a two-sample setting will create a (hopefully small) bias towards the null. In addition, because these SNPs have a weaker association, there is a greater risk that any association they do have is due to some type of confounding, which could bias estimates in either direction.

Software and Preregistration

MR analyses in this paper were run using the TwoSampleMR, MR-RAPS, MR-PRESSO, and meta R packages (9,10,15,16). The DAGs were drawn using DAGitty (17). All GWAS data was extracted from the MRC-IEU OpenGWAS platform (18).

This study was written in accordance with STROBE-MR (19), and was pre-registered at https://DOI.org/10.17605/OSF.IO/BTPH9. The quantitative assessment of whether samples were drawn from the same population was not part of our pre-registered analysis plan.

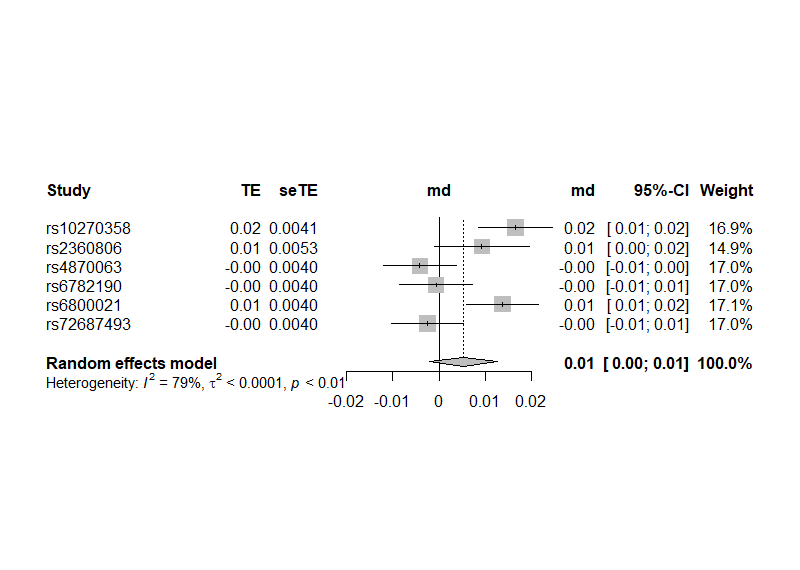

Supplementary Figure 1: Difference in SNP-outcome associations between the UKB and SSGAC when using a 5x10^-8^ p-value threshold.

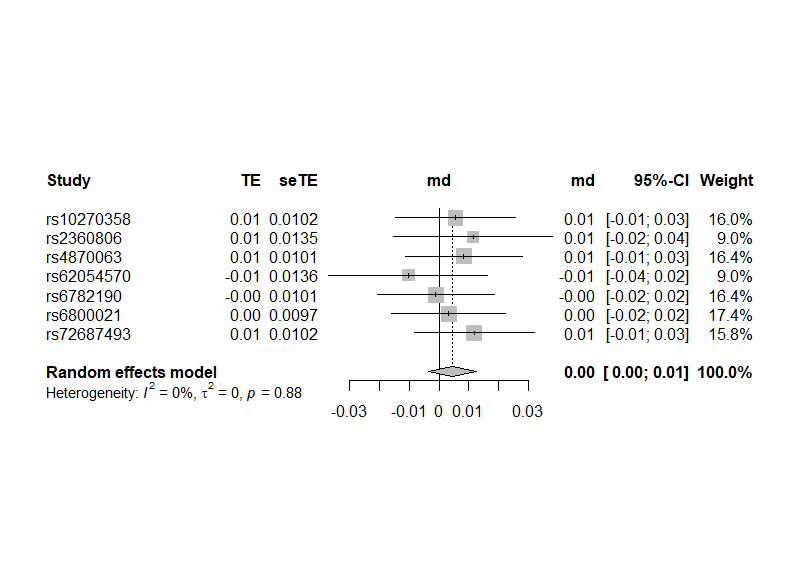

Supplementary Figure 3: Difference in SNP-outcome associations between the UKB and WFC when using a 5x10^-8^ p-value threshold.

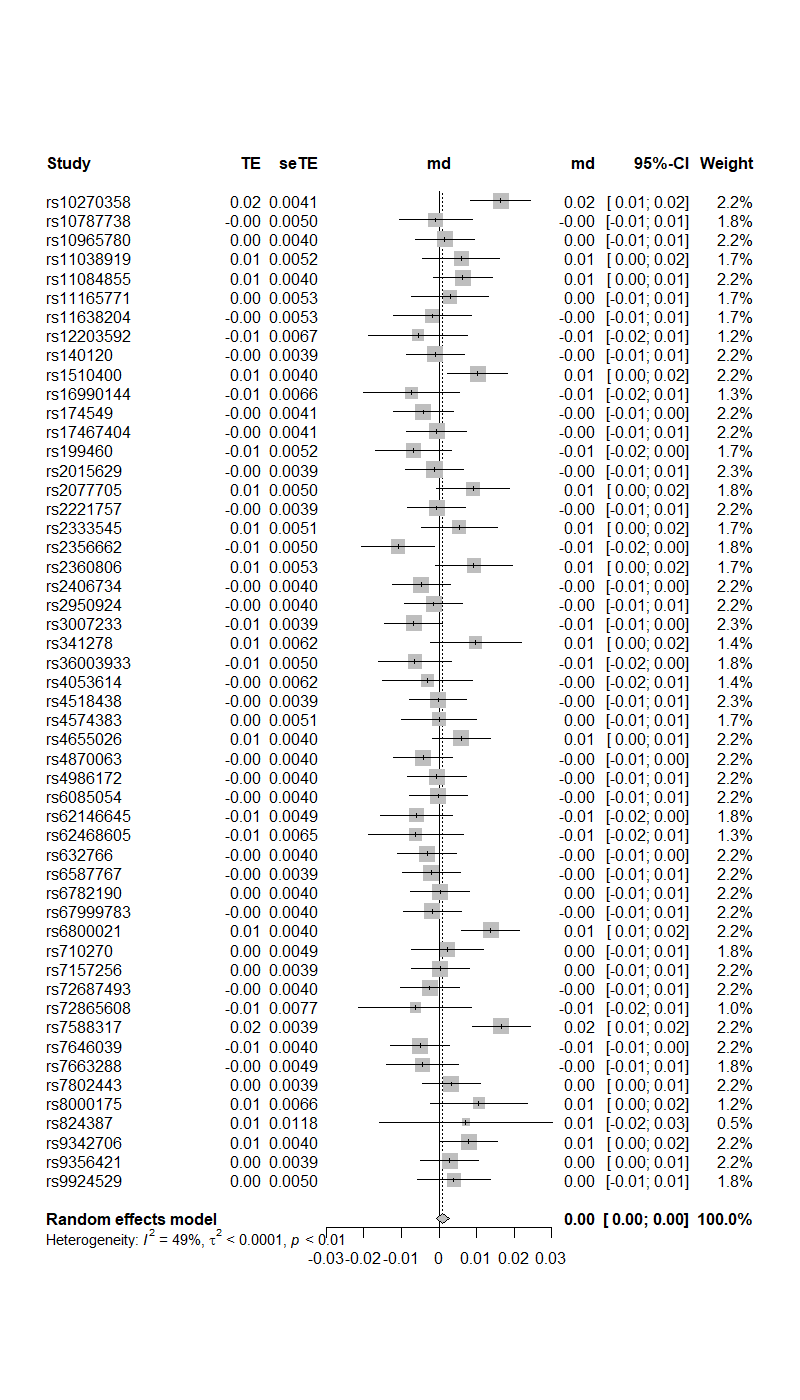
Supplementary Figure 3: Difference in SNP-outcome associations between the UKB and SSGAC when using a 5x10^-6^ p-value threshold.

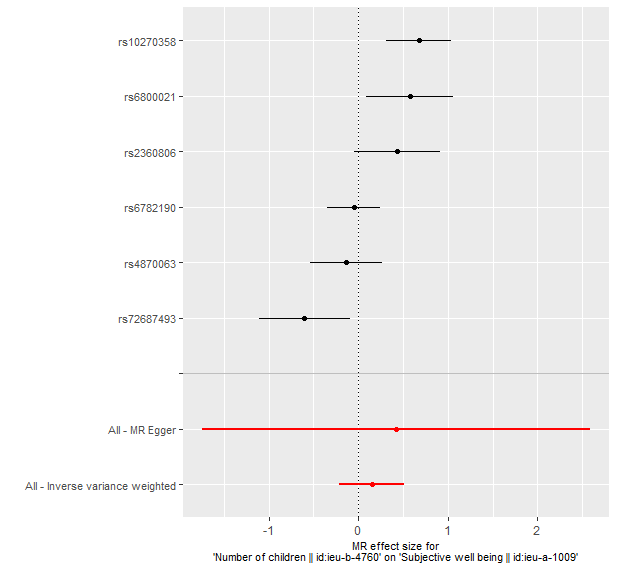

Supplementary Figure 4: Forest plot for the SNP specific Wald ratios for the primary analysis.

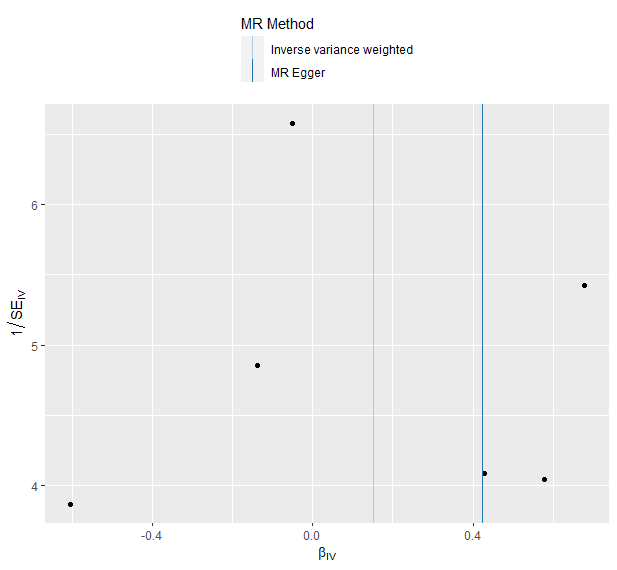

Supplementary Figure 5: Funnel plot for the primary analysis.

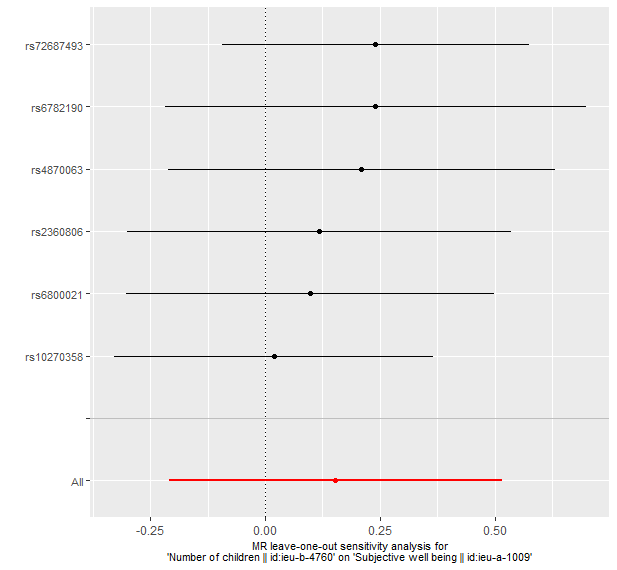

Supplementary Figure 6: Leave-one-out sensitivity analysis.
